# Does pre-infection stress increase the risk of long COVID? Longitudinal associations between adversity worries and experiences in the month prior to COVID-19 infection and the development of long COVID and specific long COVID symptoms

**DOI:** 10.1101/2022.04.06.22273444

**Authors:** Elise Paul, Daisy Fancourt

## Abstract

**Background:** Long COVID is increasingly recognised as public health burden. Demographic and infection-related characteristics have been identified as risk factors, but less research has focused on psychosocial predictors such as stress immediately preceding the index infection. Research on whether stressors predict the development of specific long COVID symptoms is also lacking.

**Methods:** Data from 1,966 UK adults who had previously been infected with COVID-19 and who took part in the UCL COVID-19 Social Study were analysed. The number of adversity experiences (e.g., job loss) and the number of worries about adversity experiences within the month prior to COVID-19 infection were used to predict the development of self-reported long COVID and the presence of three specific long COVID symptoms (difficulty with mobility, cognition, and self-care). The interaction between a three-level index of socio-economic position (SEP; with higher values indicating lower SEP) and the exposure variables in relation to long COVID status was also examined. Analyses controlled for a range of COVID-19 infection characteristics, socio-demographics, and health-related factors.

**Findings:** Odds of self-reported long COVID increased by 1.25 (95% confidence interval [CI]: 1.04 to 1.51) for each additional worry about adversity in the month prior to COVID-19 infection. Although there was no evidence for an interaction between SEP and either exposure variable, individuals in the lowest SEP group were nearly twice as likely to have developed long COVID as those in the highest SEP group (OR: 1.95; 95% CI: 1.19 to 3.19) and worries about adversity experiences remained a predictor of long COVID (OR: 1.43; 95% CI: 1.04 to 1.98). The number of worries about adversity experiences also corresponded with increased odds of certain long COVID symptoms such as difficulty with cognition (e.g., difficulty remembering or concentrating) by 1.46 (95% CI: 1.02 to 2.09) but not with mobility (e.g., walking or climbing steps) or self-care (e.g., washing all over or dressing).

**Interpretation:** Results suggest a key role of stress in the time preceding the acute COVID-19 infection for the development of long COVID and for difficulty with cognition specifically. These findings point to the importance of mitigating worries and experiences of adversities during pandemics both to reduce their psychological impact but also help reduce the societal burden of longer-term illness.

**Funding:** The Nuffield Foundation [WEL/FR-000022583], the MARCH Mental Health Network funded by the Cross-Disciplinary Mental Health Network Plus initiative supported by UK Research and Innovation [ES/S002588/1], and the Wellcome Trust [221400/Z/20/Z and 205407/Z/16/Z].

## Introduction

Though vaccines have been effective in reducing rates of severe disease and deaths from COVID-19,^1^ there are concerns that the long-term sequalae of the disease will place additional strains on the healthcare system and society. It is now recognised that a range of symptoms can persist long after the acute infection;^2^ a condition otherwise known as ‘long COVID’. This term includes both ongoing symptomatic COVID-19 (with symptoms present from 4 to 12 weeks post-onset), and post-COVID-19 syndrome (which involves the presence of symptoms more than 12 weeks post-onset).^3^ Long COVID can affect multiple organs^4–6^ and develop regardless of infection severity.^7^ Persistent symptoms typically include fatigue,^5,8–10^ breathlessness/shortness of breath (or dyspnoea), and cognitive impairments,^5,7,9–12^ also referred to as ‘brain fog’. Though definitions and measurements of long COVID vary widely and make prevalence estimates difficult to obtain, findings across studies suggest a substantial proportion of the population infected with COVID-19 will develop persisting symptoms.^9,10,12^ Long COVID often results in reduced productivity or an inability to work, and places additional strain on other aspects of one’s life. As of 2 January 2022, an estimated 2.4% of the UK population were experiencing self-reported long COVID, and nearly two-thirds (65%) said their long COVID symptoms negatively impacted their daily activities.^9^

To reduce the public health burden associated with long COVID, research has sought to identify factors predisposing its development. Most research to date has focused demographic, health, and infection-related factors, and has identified characteristics such as increasing age and female gender,^9–11,13^ higher body mass index and obesity,^6,10,14,15^ pre-existing health conditions,^6,7,9,11^ membership in an ethnic minority group,^11^ and infection severity.^6,8,10^ But less research has focused on the potential role of stress in making individuals more vulnerable to long COVID. Substantial research has documented the negative psychological toll of the pandemic globally,^16^ in particular with adverse experiences such as loss of work and income, difficulties meeting basic needs (such as food and medication), and health-related stressors and bereavement.^17^ There are a number of pathways through which such stress could predispose individuals to be more vulnerable to long COVID. First, there is a substantial literature on the relationship between both short and long-term stress and increased susceptibility to viral infections,^18–20^ as has been shown through epidemiological and intervention studies into rates of infection^18,21^ and response to vaccines.^22^ Studies exploring the pathways underlying these effects have identified that psychological stress can trigger increases in circulating markers of inflammation (e.g., interleukin (IL)-1β and tumour necrosis factor(TNF)-α)^23^ with higher inflammation in turn associated with a greater propensity for developing chronic illness.^24^ Specifically in relation to long COVID, ongoing inflammatory processes have been found to underlie the post-acute sequalae, as in other viral infections,^25–27^ and immune system dysregulation prior to infection may influence the development of long COVID.^28^ Further, research into other post-viral syndromes such as myologic encephalomyelitis/chronic fatigue syndrome (ME/CFS) has shown a heightened risk for individuals who have experienced stressful life events,^29^ and there is substantial overlap in the symptoms of long COVID and ME/CFS.^30,31^ Therefore, a relationship between stress and the development of long COVID is theoretically plausible.

Additionally, research shows that short-term stressors can be particularly detrimental for individuals with ongoing chronic stressors in their lives, as psychological and physiological coping resources are depleted, resulting in greater vulnerability to adverse effects.^32^ Lower socio-economic position (SEP) is a chronic stressor, for which the association with ill health is well established.^33^ However, findings with respect to SEP and long COVID have been mixed. Some find associations between long COVID and living in a deprived area,^9,15^. However, others find no association of high deprivation with long COVID, nor with education level,^14^ and a national registry-based Swedish study found no association between socio-economic factors (education and income) and long COVID.^34^ A large US study in Michigan found low household income to be a strong predictor of long COVID, but not after covariates were accounted for.^35^ The variation in these findings could be because stress is a mediator between SEP and increased susceptibility to long COVID. Indeed, during the COVID-19 pandemic, those from lower socio-economic backgrounds have experienced greater worries about adversities (such as loss of income or employment or challenges accessing food or medication), as well as greater experience of adversities, and these worries and experiences have had a larger effect on mental health than for individuals from higher socio-economic backgrounds.^17^ However, it remains unclear whether novel (i.e. recently occurring) stressors are a greater risk factor for long COVID amongst individuals experiencing the chronic stressor of lower SEP.

Finally, beyond a focus on risk factors for long COVID in general, it is also unknown if stress (whether chronic or shorter-term) is associated with any specific ongoing symptoms of long COVID. This is important because some long COVID symptoms may be more debilitating than others and may have different demographic and clinical risk factors.^36^ It is well known that chronic stress has a detrimental impact on cognitive functioning^37,38^ and functional impairment.^39^ There is some evidence that in select populations (e.g., caregivers for a spouse with dementia), stress may lead to cognitive, but not functional, impairments.^40^ But little research has been done on this in the context of long COVID.

Therefore, a more comprehensive understanding of whether psychosocial circumstances involving stress in the time period leading up to infection could aid in identifying early in the disease who is most likely to be negatively impacted due to ongoing symptoms. It could also have implications for the types of psychological and social support provided during pandemics. So, the first aim of this study was to explore the relationship between adversity experiences and worries about adversity experiences (stressors) and the development of self-reported long COVID. Second, we explored whether this relationship was stronger amongst individuals experiencing the chronic stressor of low SEP. Third, for people who believed themselves to have had long COVID or who had been diagnosed as such, to explore the relationship between stressors and the development of specific long COVID symptoms.

## Methods

### Study design and participants

Data were drawn from the COVID-19 Social Study; a large panel study of the psychological and social experiences of over 75,000 adults (aged 18+) in the UK during the COVID-19 pandemic. The study commenced on 21 March 2020 and involves online data collection from participants for the duration of the COVID-19 pandemic in the UK. Data were initially collected weekly (from study commencement through August 2020), then monthly thereafter. The study is not random and therefore is not representative of the UK population. But it does contain a well-stratified sample that was recruited using three primary approaches. First, convenience sampling was used, including promoting the study through existing networks and mailing lists (including large databases of adults who had previously consented to be involved in health research across the UK), print and digital media coverage, and social media. Second, more targeted recruitment was undertaken focusing on groups who were anticipated to be less likely to take part in the research via our first strategy, including (i) individuals from a low-income background, (ii) individuals with no or few educational qualifications, and (iii) individuals who were unemployed. Third, the study was promoted via partnerships with third sector organisations to vulnerable groups, including adults with pre-existing mental health conditions, older adults, carers, and people experiencing domestic violence or abuse. The study was approved by the UCL Research Ethics Committee [12467/005] and all participants gave informed consent. Participants were not compensated for participation (https://osf.io/jm8ra/).

We included participants who met the following criteria. First, participants were included if they said in November 2021 that they had at some prior point been infected with COVID-19. Second, they had to have provided a date for their COVID-19 infection which was no earlier than 7 April 2020, and at least five weeks^9,10^ before follow-up (survey distributed the week of 22 November 2021, with participant responses to 10 January 2022). 7 April 2020 was chosen as we were interested in adversity experiences and worries about adversity within the month prior (range 1-8 weeks) to COVID-19 infection, and the collection of all individual items comprising these variables commenced 30 March 2020. Five weeks was chosen as the minimum time period as many studies on long COVID apply a threshold of “more than four weeks of symptoms” to be experienced for the term long COVID to be applied and our outcome measures asked about symptoms in the past week.^9,10^ Third, participants who had had COVID-19 only once were included; participants who reported more than one infection were excluded. Fourth, participants had to have taken part in the survey at least once in the month prior (range 1-8 weeks) to the date of their infection. Fifth, they had to have non-missing data on long COVID outcome variables (presence/absence and specific long COVID symptoms [for people with long COVID]) and study variables required to calculate statistical weights (gender, age, ethnicity, country, and education). The final analytic sample comprised 1,966. See Figure 1 for a flow chart of the number of participants excluded at each step.

**Figure 1.**
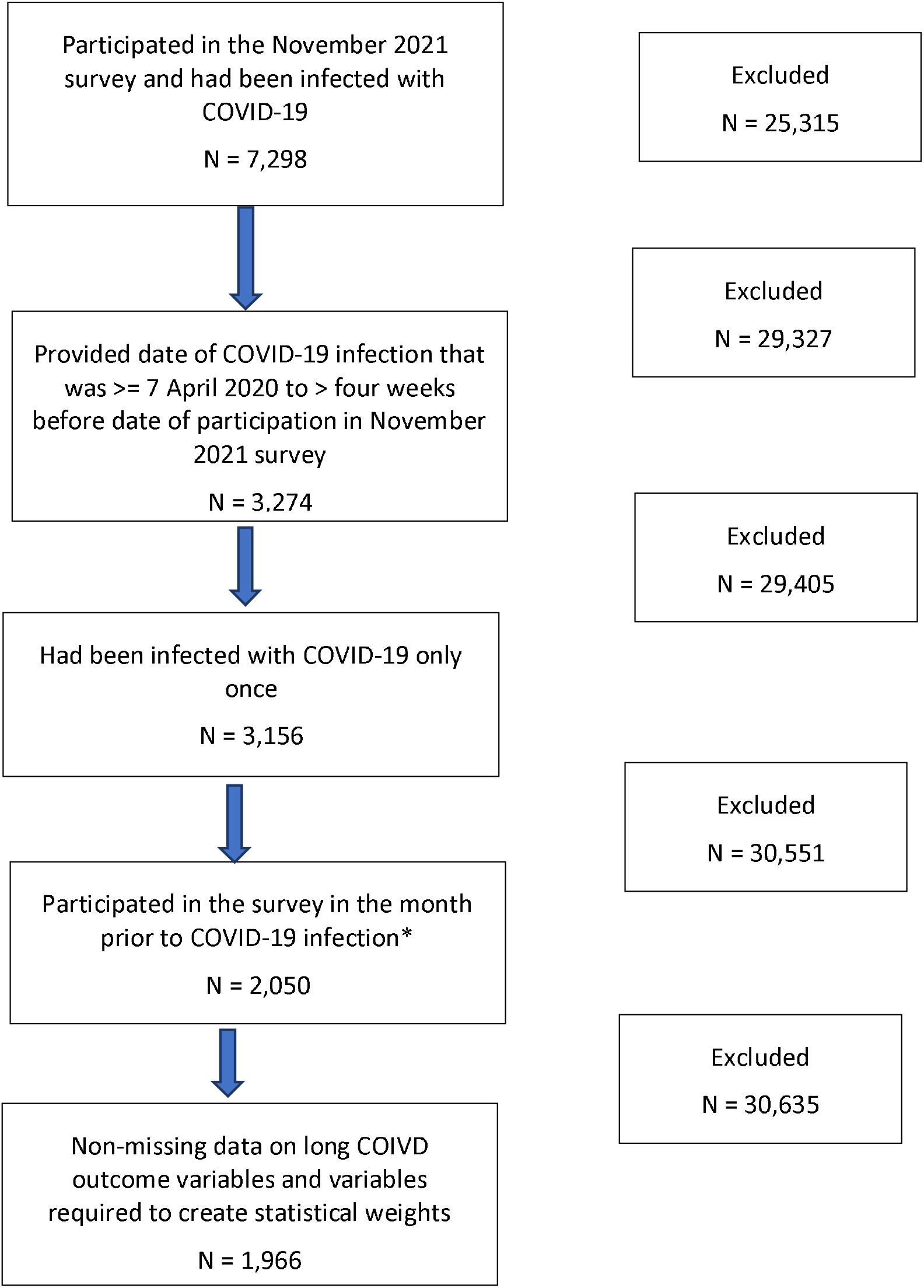
Flow chart of sample selection. *Starting with the week before infection, up to 8 weeks prior to infection, when earlier weeks were missing

We used multiple imputation by chained equations to generate 50 imputed datasets for participants who met the above outlined study inclusion criteria but had missing data on other study variables. Proportions of missing data ranged from 0.81% for the severity of COVID-19 infection variable to 7.94% for household income (Supplemental Table S1). Imputation models included all study variables, as well as additional auxiliary variables collected at baseline (e.g., home ownership status, and depressive symptoms). Substantive results using cases without any missing data (i.e., complete case analysis) and the imputed sample were similar (Supplemental Tables S2-S6). See Supplemental Table S7 for a comparison of excluded and included participants on study variables.

## Measures

### Outcome variables

For our first research question, the presence or absence of long COVID was measured with response to the question: “Do you consider yourself to have (or have had) Long Covid?”. The four response options (given in Supplemental Table S8) were categorised into (i) no and (ii) yes (formally diagnosed or suspected). Sensitivity analyses were carried out to test whether results were consistent when including participants who were “unsure” about whether they had had long COVID within the case group. Due to the self-reported nature of our long COVID outcome variable, an additional sensitivity analysis was also carried out with participants reporting COVID-19 symptoms four weeks or more as the case group and compared to those whose symptoms lasted less than four weeks. See Supplemental Table S8 for question wording.

For our second research question, we restricted our sample just to people with long COVID to look at the presence of three specific long COVID symptoms. These three variables were operationalised from questions assessing the extent to which participants had difficulty with (i) mobility, (ii) cognition, and (iii) self-care (Supplemental Table S8). Response options were on a four-point scale ranging from 0 “no difficulty at all” to “unable to do” and treated as binary (present vs absent) in analyses due to low numbers within response categories.

### Predictor variables

#### Adversity experiences and worry about adversity experiences in the month before COVID-19 infection

Two variables were created to represent the total number of adversity experiences and the total number of worries about adversity in the month (range 1-8 weeks) prior to each participant’s COVID-19 infection. Data on adversity experiences and worry about adversity experiences in the month prior to infection were prioritised in order of proximity to the infection. For example, if data collected one week prior to the COVID-19 infection were not available, data two weeks prior were used, up to eight weeks before the infection. The majority of the sample (91.71%) had data within four weeks prior to the infection. See Supplemental Table S9 for the distribution of the number of weeks prior to infection for which participants provided data.

The categories of adversity (10 items) and worries (11 items) were comparable: (i) financial, (ii) accessing essentials, (iii) bereavement or other social/relationship issues, and (iv) abuse/safety and security. See Supplemental Table S8 for a full listing of items. Each of the binary (absent vs present) adversity and worry about adversity items were summed to create the total number of adversity experiences and worries about adversity. Scores were then standardised to facilitate comparability of resulting odds ratios in logistic regression models.

#### Socio-economic position

We constructed a three-level index of socio-economic position (SEP) using five variables collected at baseline participant’s first round of data collection: annual household income of less than <£30,000, highest educational qualification up to GCSE (General Certificate of Secondary Education) [equivalent to education to age 16]), low unemployed, renting or living rent-free (vs own outright or own with mortgage), and household overcrowding (>1 persons per room). The index was collapsed into 0, 1 and 2+ indications of low SEP to attain adequate sample sizes for each category.

### Covariates

#### COVID-19 infection variables

The severity of COVID-19 infection was assessed with the study developed question: “How severe were your symptoms in the first 1-2 weeks?”. Response options were categorised into (i) asymptomatic, (ii) mild (experienced symptoms but was able to carry on with daily activities), (iii) moderate (experienced symptoms and had to rest in bed), and (iv) severe (participant was hospitalised).

A variable to indicate which strain of the virus was dominant in the UK^41^ at the time of infection was included and coded as (0) the original COVID-19 variant (31 January to 31 October 2020, (1) Alpha (1 November 2020 to 30 June 2021), (2) Delta (1 July 2021 to 30 November 2021), and (3) Omicron (1 December 2021 onwards).

#### Socio-demographics

Socio-demographic factors were collected at baseline and included gender (male vs female), age group (60+, 45-59, 30-44, and 18-29) ethnicity (white vs ethnic minority groups [i.e., Asian/Asian British, Black/Black British. See Supplemental Table S8 for a full listing of response options]), government’s identified key worker status (not a key worker vs key worker), living arrangement (living alone vs living with others but not including children vs living with others, including children), and area of dwelling (urban [city, large town, small town] vs rural [village, hamlet, isolated dwelling]). Key workers included people with jobs deemed essential during the pandemic (e.g., health and social care, education, and childcare) and who were required to leave home to carry out this work during lockdowns.

#### Health-related factors

We also included two health-related factors measured at baseline. Participants reported whether they had received a clinical diagnosis of a mental health condition clinically-diagnosed depression, clinically-diagnosed anxiety, or another clinically-diagnosed mental health problem) or chronic physical health condition (high blood pressure, diabetes, heart disease, lung disease (e.g. asthma or COPD), cancer, another clinically-diagnosed chronic physical health condition, a disability that affects the ability to leave the house, any other disability, and pregnancy). Responses were used to create two binary variables to indicate the presence or absence of pre-existing physical and mental health conditions.

### Statistical analysis

Three sets of analyses were carried out. First, binary logistic regression models were fitted to examine associations of predictor variables (adversity experiences and worries about adversity) and the development of long COVID. Second, we fitted an interaction term between experiences and worries and the low SEP index. Third, for individuals with long COVID, binary logistic regression models were fitted to examine associations between predictor variables and the presence of each of the three specific long COVID symptoms (difficulty with mobility, cognition, and self-care).

For all sets of analyses, Model 1 included only adversity experiences and worries about adversity experiences in the same model (plus the interaction terms with SEP for analysis (2)), Model 2 additionally adjusted for COVID-19 infection variables, Model 3 additionally adjusted for socio-demographic characteristics, and Model 4 additionally adjusted for health-related factors. Robust standard errors were used in all analyses. Coefficients from the binary logistic regressions were exponentiated and presented as odds ratios (OR), along with corresponding 95% confidence intervals (CI).

To account for the non-random nature of the sample and increase representativeness of the UK general population, all data were weighted to the proportions of gender, age, ethnicity, country, and education obtained from the Office for National Statistics.^42^ A multivariate reweighting method was implemented using the Stata user written command ‘ebalance’.^43^ Further details are shown in the User Guide (https://osf.io/jm8ra/). Analyses were conducted using Stata version 16.^44^

## Results

### Sample characteristics

Unweighted and weighted characteristics of the study sample are presented in Supplemental Table S10. Before weighting, the sample was disproportionately female, White, had higher levels of education and household income, and of older age. However, differences in other study variables were minimal. 20.10% of the weighted sample reported that they had either been diagnosed with (3.76%) or believed they had had (16.34%) long COVID, whilst 12.41% were unsure. A number of different socio-demographic groups were more likely to develop long COVID (Table 1): adults aged 45-59, people from ethnic minority groups, those with lower levels of education and household income, people living with children, people who live in crowded accommodation, those who live in a rural area, who had had moderate or severe COVID-19, and people with a mental or physical health condition. Individuals with long COVID and who were unsure whether they had long COVID were also more likely to be in the lowest SEP category (2+) (long COVID group: 59.61%; unsure group: 52.72%) compared to the group without long COVID (42.16%). The proportion with long COVID who had been infected with COVID-19 at the time the Alpha strain (46.97%) was dominant in the UK was higher than that of the Delta variant (33.42%), and no participants had been infected at the time Omicron was dominant (Table 1). Difficulty with cognition was the most often reported specific long COVID symptom in the long COVID group and amongst those who were unsure about having had long COVID (long COVID group: 62.07%; unsure group: 41.16%), followed by difficulty with mobility (long COVID group: 53.52%; unsure group: 28.74%). Most (83.17%) individuals with long COVID who reported difficulty with self-care were in the lowest SEP group, followed by those who had difficulty with mobility (68.84%) and difficulty with cognition (56.52%) (Table 2).

**Table 1.**
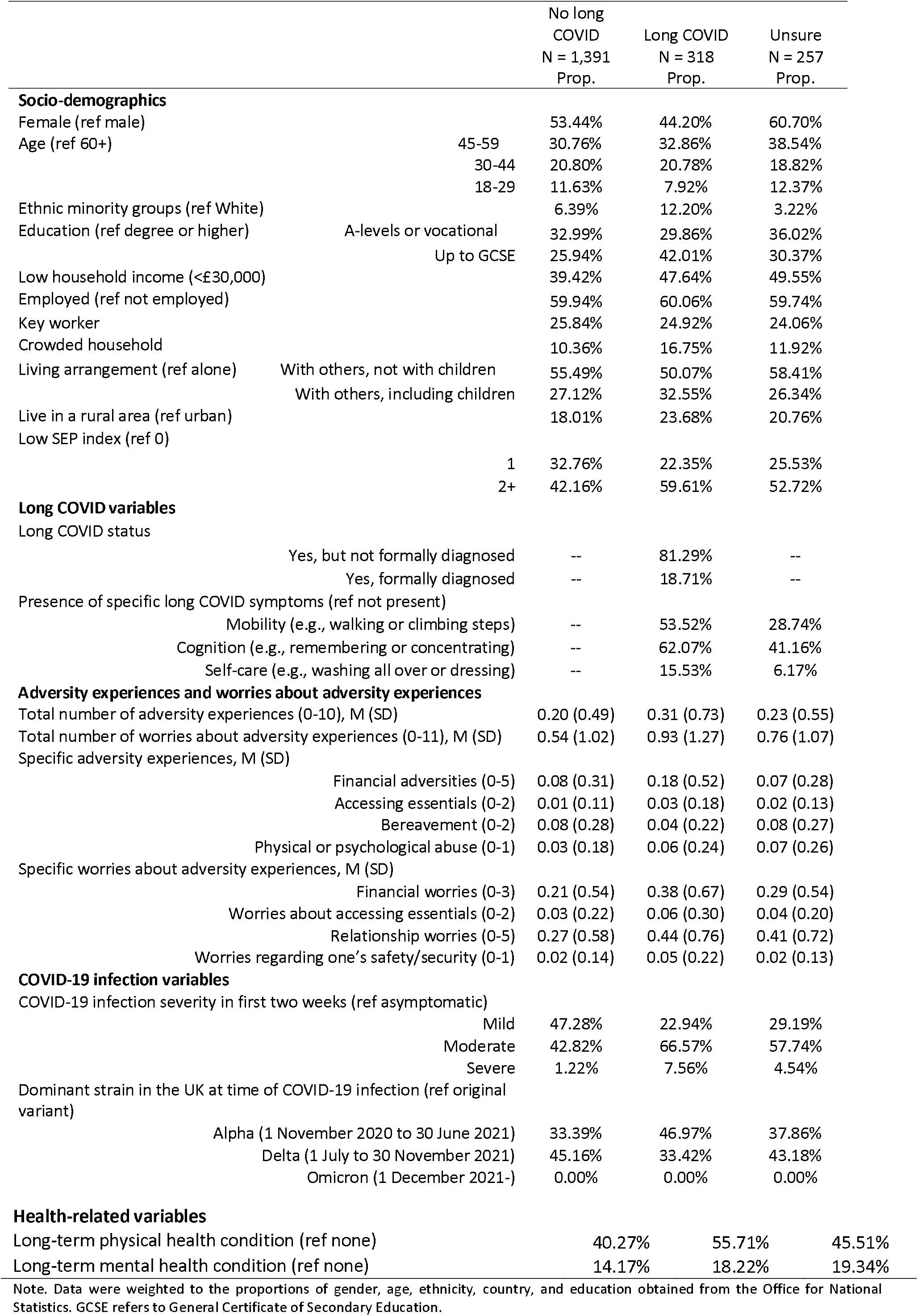
Sample characteristics by long COVID status, weighted (N = 1,966)

|  |  | No long<br>COVID<br>N = 1,391<br>Prop. | Long COVID<br>N = 318<br>Prop. | Unsure<br>N = 257<br>Prop. |
| --- | --- | --- | --- | --- |
| **Socio-demographics** |  |  |  |  |
| Female (ref male) |  | 53.44% | 44.20% | 60.70% |
| Age (ref 60+) | 45-59 | 30.76% | 32.86% | 38.54% |
|  | 30-44 | 20.80% | 20.78% | 18.82% |
|  | 18-29 | 11.63% | 7.92% | 12.37% |
| Ethnic minority groups (ref White) |  | 6.39% | 12.20% | 3.22% |
| Education (ref degree or higher) | A-levels or vocational | 32.99% | 29.86% | 36.02% |
|  | Up to GCSE | 25.94% | 42.01% | 30.37% |
| Low household income (<£30,000) |  | 39.42% | 47.64% | 49.55% |
| Employed (ref not employed) |  | 59.94% | 60.06% | 59.74% |
| Key worker |  | 25.84% | 24.92% | 24.06% |
| Crowded household |  | 10.36% | 16.75% | 11.92% |
| Living arrangement (ref alone) | With others, not with children | 55.49% | 50.07% | 58.41% |
|  | With others, including children | 27.12% | 32.55% | 26.34% |
| Live in a rural area (ref urban) |  | 18.01% | 23.68% | 20.76% |
| Low SEP index (ref 0) | 1 | 32.76% | 22.35% | 25.53% |
|  | 2+ | 42.16% | 59.61% | 52.72% |
| **Long COVID variables** |  |  |  |  |
| Long COVID status | Yes, but not formally diagnosed | -- | 81.29% | -- |
|  | Yes, formally diagnosed | -- | 18.71% | -- |
| Presence of specific long COVID symptoms (ref not present) |  |  |  |  |
|  | Mobility (e.g., walking or climbing steps) | -- | 53.52% | 28.74% |
|  | Cognition (e.g., remembering or concentrating) | -- | 62.07% | 41.16% |
|  | Self-care (e.g., washing all over or dressing) | -- | 15.53% | 6.17% |
| **Adversity experiences and worries about adversity experiences** |  |  |  |  |
| Total number of adversity experiences (0-10), M (SD) |  | 0.20 (0.49) | 0.31 (0.73) | 0.23 (0.55) |
| Total number of worries about adversity experiences (0-11), M (SD) |  | 0.54 (1.02) | 0.93 (1.27) | 0.76 (1.07) |
| Specific adversity experiences, M (SD) |  |  |  |  |
|  | Financial adversities (0-5) | 0.08 (0.31) | 0.18 (0.52) | 0.07 (0.28) |
|  | Accessing essentials (0-2) | 0.01 (0.11) | 0.03 (0.18) | 0.02 (0.13) |
|  | Bereavement (0-2) | 0.08 (0.28) | 0.04 (0.22) | 0.08 (0.27) |
|  | Physical or psychological abuse (0-1) | 0.03 (0.18) | 0.06 (0.24) | 0.07 (0.26) |
| Specific worries about adversity experiences, M (SD) |  |  |  |  |
|  | Financial worries (0-3) | 0.21 (0.54) | 0.38 (0.67) | 0.29 (0.54) |
|  | Worries about accessing essentials (0-2) | 0.03 (0.22) | 0.06 (0.30) | 0.04 (0.20) |
|  | Relationship worries (0-5) | 0.27 (0.58) | 0.44 (0.76) | 0.41 (0.72) |
|  | Worries regarding one's safety/security (0-1) | 0.02 (0.14) | 0.05 (0.22) | 0.02 (0.13) |
| **COVID-19 infection variables** |  |  |  |  |
| COVID-19 infection severity in first two weeks (ref asymptomatic) |  |  |  |  |
|  | Mild | 47.28% | 22.94% | 29.19% |
|  | Moderate | 42.82% | 66.57% | 57.74% |
|  | Severe | 1.22% | 7.56% | 4.54% |
| Dominant strain in the UK at time of COVID-19 infection (ref original variant) |  |  |  |  |
|  | Alpha (1 November 2020 to 30 June 2021) | 33.39% | 46.97% | 37.86% |
|  | Delta (1 July to 30 November 2021) | 45.16% | 33.42% | 43.18% |
|  | Omicron (1 December 2021-) | 0.00% | 0.00% | 0.00% |

**Health-related variables**
|  |  |  |  |
| --- | --- | --- | --- |
| Long-term physical health condition (ref none) | 40.27% | 55.71% | 45.51% |
| Long-term mental health condition (ref none) | 14.17% | 18.22% | 19.34% |

Note. Data were weighted to the proportions of gender, age, ethnicity, country, and education obtained from the Office for National Statistics. GCSE refers to General Certificate of Secondary Education.

**Table 2.** Characteristics of participants with long COVID by specific long COVID symptoms (N = 318), weighted

|  |  | Difficulty<br>with<br>mobility<br>N = 172<br>Prop. | Difficulty<br>with<br>cognition<br>N = 217<br>Prop. | Difficulty<br>with self-care<br>N = 50<br>Prop. |
| --- | --- | --- | --- | --- |
| **Socio-demographics** |  |  |  |  |
| Female (ref male) |  | 40.51% | 51.56% | 46.24% |
| Age (ref 60+) | 45-59 | 31.94% | 36.31% | 31.42% |
|  | 30-44 | 13.20% | 22.85% | 6.73% |
|  | 18-29 | 1.52% | 1.17% | 0.00% |
| Ethnic minority groups (ref White) |  | 5.11% | 14.23% | 8.55% |
| Education (ref degree or higher) | A-levels or vocational | 25.49% | 26.39% | 28.73% |
|  | Up to GCSE | 49.68% | 48.35% | 48.03% |
| Low household income (<£30,000) |  | 62.12% | 47.10% | 73.15% |
| Employed (ref not employed) |  | 51.94% | 65.65% | 30.27% |
| Key worker |  | 16.95% | 27.92% | 14.65% |
| Crowded household |  | 13.43% | 14.62% | 14.24% |
| Living arrangement (ref alone) | With others, not with children | 52.78% | 49.16% | 48.91% |
|  | With others, including children | 25.26% | 32.71% | 28.96% |
| Live in a rural area (ref urban) |  | 23.08% | 23.11% | 26.10% |
| Low SEP index (ref 0) |  |  |  |  |
|  | 1 | 15.88% | 24.48% | 8.86% |
|  | 2+ | 68.84% | 56.52% | 83.17% |
| **Long COVID variables** |  |  |  |  |
| Long COVID status |  |  |  |  |
|  | Yes, but not formally diagnosed | 73.50% | 76.47% | 60.96% |
|  | Yes, formally diagnosed | 26.50% | 23.53% | 39.04% |
| Presence of specific long COVID symptoms (ref not present) |  |  |  |  |
|  | Mobility (e.g., walking or climbing steps) | -- | 57.87% | 97.76% |
|  | Cognition (e.g., remembering or concentrating) | 67.12% | -- | 85.54% |
|  | Self-care (e.g., washing all over or dressing) | 28.36% | 21.40% | -- |
| **Adversity experiences and worries about adversity experiences** |  |  |  |  |
| Total number of adversity experiences (0-10), M (SD) |  | 0.39 (0.82) | 0.41 (0.85) | 0.55 (0.93) |
| Total number of worries about adversity experiences (0-11), M (SD) |  | 1.05 (1.39) | 1.07 (1.44) | 1.64 (1.90) |
| Specific adversity experiences, M (SD) |  |  |  |  |
|  | Financial adversities (0-5) | 0.20 (0.54) | 0.25 (0.62) | 0.23 (0.55) |
|  | Accessing essentials (0-2) | 0.05 (0.22) | 0.05 (0.22) | 0.08 (0.30) |
|  | Bereavement (0-2) | 0.04 (0.22) | 0.04 (0.24) | 0.03 (0.20) |
|  | Physical or psychological abuse (0-1) | 0.10 (0.30) | 0.07 (0.26) | 0.20 (0.41) |
| Specific worries about adversity experiences, M (SD) |  |  |  |  |
|  | Financial worries (0-3) | 1.02 (1.42) | 1.17 (1.51) | 1.18 (1.37) |
|  | Worries about accessing essentials (0-2) | 0.28 (0.79) | 0.28 (0.82) | 0.61 (1.25) |
|  | Relationship worries (0-5) | 1.70 (1.76) | 1.63 (1.70) | 2.26 (2.34) |
|  | Worries regarding one's safety/security (0-1) | 0.19 (0.53) | 0.14 (0.46) | 0.40 (0.76) |
| **COVID-19 infection variables** |  |  |  |  |
| COVID-19 infection severity in first two weeks (ref asymptomatic) |  |  |  |  |
|  | Mild | 18.61% | 16.48% | 3.77% |
|  | Moderate | 65.19% | 72.11% | 75.32% |
|  | Severe | 11.86% | 8.37% | 14.03% |
| Dominant strain in the UK at time of COVID-19 infection (ref original variant) |  |  |  |  |
|  | Alpha (1 November 2020 to 30 June 2021) | 46.93% | 49.96% | 38.66% |
|  | Delta (1 July to 30 November 2021) | 34.21% | 36.03% | 38.80% |
|  | Omicron (1 December 2021-) | 0.00% | 0.00% | 0.00% |
| **Health-related variables** |  |  |  |  |
| Long-term physical health condition (ref none) |  | 65.80% | 57.77% | 79.76% |
| Long-term mental health condition (ref none) |  | 17.15% | 19.72% | 24.19% |
| Note. Data were weighted to the proportions of gender, age, ethnicity, country, and education obtained from the Office for National Statistics. GCSE refers to General Certificate of Secondary Education. |  |  |  |  |

### Adversity experiences and worries

The average numbers of adversity experiences and worries about adversity experiences in the month prior to the COVID-19 infection were higher in the long COVID and unsure groups, compared to the group that did not develop long COVID. The average numbers of financial adversities, adversities related to accessing essentials, and psychological/physical abuse were twice as high in the long COVID group compared to the group without long COVID (Table 1). Worries about all of the specific adversity experiences were also higher in the long COVID group. In the group with long COVID, individuals who reported difficulties with self-care had experienced more adversities in the month before infection with COVID-19 and were also more worried about adversities during this time (Table 2). They were also twice as likely as individuals with the other two long COVID symptoms to have been abused. Individuals with cognition difficulties were more likely to be female, of older age, from an ethnic minority group, a key worker, employed, and over half (57.87%) also had difficulty with mobility (Table 2).

Results from binary logistic regressions predicting the of long COVID are presented in Table 3. Worries about adversity experiences were associated with an increased likelihood of developing long COVID (odds ratio [OR]: 1.25; 95% confidence interval [CI]: 1.04 to 1.51), but adversity experiences were not (OR: 1.07; 95% CI: 0.91 to 1.26) in the fully adjusted model.

**Table 3.** Logistic regressions predicting self-reported long COVID from adversity experiences and worries about adversity experiences (N = 1,709), weighted

|  | Self-reported long COVID |  |  |  |  |  |
| --- | --- | --- | --- | --- | --- | --- |
|  | OR | SE | T | P | 95% CI | 95% CI |
| **Model 1** |  |  |  |  |  |  |
| Total number of adversity experiences | 1.07 | 0.08 | 0.85 | 0.39 | 0.92 | 1.25 |
| Total number of worries about adversity experiences | **1.32** | **0.11** | **3.38** | **<0.001** | **1.12** | **1.56** |
| **Model 2** |  |  |  |  |  |  |
| Total number of adversity experiences | 1.04 | 0.08 | 0.52 | 0.60 | 0.89 | 1.22 |
| Total number of worries about adversity experiences | **1.28** | **0.11** | **2.97** | **<0.001** | **1.09** | **1.51** |
| **Model 3** |  |  |  |  |  |  |
| Total number of adversity experiences | 1.06 | 0.09 | 0.75 | 0.45 | 0.90 | 1.25 |
| Total number of worries about adversity experiences | **1.26** | **0.12** | **2.54** | **0.01** | **1.06** | **1.51** |
| **Model 4** |  |  |  |  |  |  |
| Total number of adversity experiences | 1.07 | 0.09 | 0.80 | 0.42 | 0.91 | 1.26 |
| Total number of worries about adversity experiences | **1.25** | **0.12** | **2.38** | **0.02** | **1.04** | **1.51** |

Note. Data were weighted to the proportions of gender, age, ethnicity, country, and education obtained from the Office for National Statistics. Model 1 included only adversity experiences and worries about adversity experiences (in the same model), Model 2 additionally adjusted for COVID-19 infection variables, Model 3 additionally adjusted for socio-demographic characteristics, and Model 4 additionally adjusted for health-related factors.

### Adversities and SEP

In the fully adjusted model, which included the interaction between the SEP index and the two exposure variables, worries about adversity experiences was associated with long COVID (OR: 1.43; 95% CI: 1.04 to 1.98) (Table 4). Additionally, people in the lowest SEP category were nearly twice as likely to have developed long COVID than those in the highest SEP group (OR: 1.95; 95% CI: 1.19 to 2.19). However, there was no evidence for an interaction between the SEP index and either exposure variable.

**Table 4.** Logistic regressions predicting self-reported long COVID from adversity experiences and worries about adversity experiences in interaction with SEP (N = 1,709), weighted

|  |  | Self-reported long COVID |  |  |  |  |  |
| --- | --- | --- | --- | --- | --- | --- | --- |
|  |  | OR | SE | T | P | 95% CI | 95% CI |
| **Model 1** |  |  |  |  |  |  |  |
| Low SEP index (ref 0) |  |  |  |  |  |  |  |
|  | 1 | 0.94 | 0.24 | -0.25 | 0.81 | 0.58 | 1.54 |
|  | 2+ | **1.84** | **0.38** | **2.97** | **<0.001** | **1.23** | **2.75** |
| Total number of adversity experiences |  | 1.15 | 0.16 | 0.98 | 0.33 | 0.87 | 1.51 |
| Interaction with SEP |  |  |  |  |  |  |  |
|  | 1 | 0.92 | 0.21 | -0.35 | 0.72 | 0.60 | 1.43 |
|  | 2+ | 0.92 | 0.16 | -0.51 | 0.61 | 0.65 | 1.29 |
| Total number of worries about adversity experiences |  | **1.33** | **0.19** | **2.03** | **0.04** | **1.01** | **1.76** |
| Interaction with SEP |  |  |  |  |  |  |  |
|  | 1 | 0.84 | 0.16 | -0.92 | 0.36 | 0.57 | 1.22 |
|  | 2+ | 1.03 | 0.19 | 0.16 | 0.88 | 0.72 | 1.48 |
| **Model 2** |  |  |  |  |  |  |  |
| Low SEP index (ref 0) |  |  |  |  |  |  |  |
|  | 1 | 0.92 | 0.23 | -0.34 | 0.73 | 0.56 | 1.49 |
|  | 2+ | **1.85** | **0.40** | **2.87** | **<0.001** | **1.22** | **2.82** |
| Total number of adversity experiences |  | 1.14 | 0.17 | 0.87 | 0.38 | 0.85 | 1.54 |
| Interaction with SEP |  |  |  |  |  |  |  |
|  | 1 | 0.89 | 0.21 | -0.52 | 0.61 | 0.56 | 1.40 |
|  | 2+ | 0.90 | 0.17 | -0.54 | 0.59 | 0.63 | 1.30 |
| Total number of worries about adversity experiences |  | **1.38** | **0.20** | **2.21** | **0.03** | **1.04** | **1.84** |
| Interaction with SEP |  |  |  |  |  |  |  |
|  | 1 | 0.74 | 0.16 | -1.42 | 0.15 | 0.48 | 1.12 |
|  | 2+ | 0.96 | 0.19 | -0.20 | 0.84 | 0.66 | 1.40 |
| **Model 3** |  |  |  |  |  |  |  |
| Low SEP index (ref 0) |  |  |  |  |  |  |  |
|  | 1 | 0.86 | 0.21 | -0.62 | 0.53 | 0.53 | 1.39 |
|  | 2+ | **2.04** | **0.51** | **2.87** | **<0.001** | **1.25** | **3.33** |
| Total number of adversity experiences |  | 1.12 | 0.19 | 0.65 | 0.52 | 0.80 | 1.55 |
| Interaction with SEP |  |  |  |  |  |  |  |
|  | 1 | 0.92 | 0.23 | -0.35 | 0.73 | 0.56 | 1.50 |
|  | 2+ | 0.96 | 0.19 | -0.20 | 0.84 | 0.65 | 1.42 |
| Total number of worries about adversity experiences |  | **1.43** | **0.23** | **2.24** | **0.02** | **1.05** | **1.95** |
| Interaction with SEP |  |  |  |  |  |  |  |
|  | 1 | 0.72 | 0.16 | -1.42 | 0.15 | 0.46 | 1.13 |
|  | 2+ | 0.90 | 0.19 | -0.49 | 0.63 | 0.60 | 1.36 |
| **Model 4** |  |  |  |  |  |  |  |
| Low SEP index (ref 0) |  |  |  |  |  |  |  |
|  | 1 | 0.84 | 0.21 | -0.69 | 0.49 | 0.51 | 1.38 |
|  | 2+ | **1.95** | **0.49** | **2.65** | **0.01** | **1.19** | **3.19** |
| Total number of adversity experiences |  | 1.12 | 0.19 | 0.67 | 0.50 | 0.81 | 1.55 |
| Interaction with SEP |  |  |  |  |  |  |  |
|  | 1 | 0.93 | 0.23 | -0.31 | 0.76 | 0.57 | 1.50 |
|  | 2+ | 0.96 | 0.19 | -0.20 | 0.84 | 0.65 | 1.42 |
| Total number of worries about adversity experiences |  | **1.43** | **0.23** | **2.21** | **0.03** | **1.04** | **1.98** |
| Interaction with SEP |  |  |  |  |  |  |  |
|  | 1 | 0.71 | 0.17 | -1.46 | 0.14 | 0.45 | 1.12 |
|  | 2+ | 0.90 | 0.19 | -0.52 | 0.60 | 0.59 | 1.36 |

Note. Data were weighted to the proportions of gender, age, ethnicity, country, and education obtained from the Office for National Statistics. Model 1 included only adversity experiences and worries about adversity experiences (in the same model), Model 2 additionally adjusted for COVID-19 infection variables, Model 3 additionally adjusted for socio-demographic characteristics, and Model 4 additionally adjusted for health-related factors.

### Specific long COVID symptoms

When looking at predictors of specific long COVID symptoms, only worries about adversities and not adversity experiences, were associated with difficulty with cognition and self-care, but not with mobility (Tables 5-7). In the full model, worries about adversity experiences remained a significant predictor of difficulty with cognition (OR: 1.46; 95% CI: 1.02 to 2.09; Table 6), but not with self-care (OR: 1.30; 95% CI: 0.93 to 1.81; Table 7).

**Table 5.** Logistic regressions predicting the development of difficulty with mobility from adversity experiences and worries about adversity experiences (N = 318), weighted

|  | Difficulty with mobility |  |  |  |  |  |
| --- | --- | --- | --- | --- | --- | --- |
|  | OR | SE | T | P | 95% CI | 95% CI |
| **Model 1** |  |  |  |  |  |  |
| Total number of adversity experiences | 1.18 | 0.16 | 1.25 | 0.21 | 0.91 | 1.54 |
| Total number of worries about adversity experiences | 1.09 | 0.17 | 0.59 | 0.55 | 0.81 | 1.47 |
| **Model 2** |  |  |  |  |  |  |
| Total number of adversity experiences | 1.20 | 0.16 | 1.35 | 0.18 | 0.92 | 1.56 |
| Total number of worries about adversity experiences | 1.09 | 0.16 | 0.56 | 0.57 | 0.81 | 1.46 |
| **Model 3** |  |  |  |  |  |  |
| Total number of adversity experiences | 1.30 | 0.19 | 1.83 | 0.07 | 0.98 | 1.73 |
| Total number of worries about adversity experiences | 1.02 | 0.16 | 0.15 | 0.88 | 0.76 | 1.39 |
| **Model 4** |  |  |  |  |  |  |
| Total number of adversity experiences | 1.33 | 0.21 | 1.79 | 0.07 | 0.97 | 1.82 |
| Total number of worries about adversity experiences | 1.05 | 0.17 | 0.27 | 0.79 | 0.76 | 1.44 |

Note. Data were weighted to the proportions of gender, age, ethnicity, country, and education obtained from the Office for National Statistics. Model 1 included only adversity experiences and worries about adversity experiences (in the same model), Model 2 additionally adjusted for COVID-19 infection variables, Model 3 additionally adjusted for socio-demographic characteristics, and Model 4 additionally adjusted for health-related factors.

**Table 6.** Logistic regressions predicting the development of difficulty with cognition from adversity experiences and worries about adversity experiences (N = 318), weighted

|  | Difficulty with cognition |  |  |  |  |  |
| --- | --- | --- | --- | --- | --- | --- |
|  | OR | SE | T | P | 95% CI | 95% CI |
| **Model 1** |  |  |  |  |  |  |
| Total number of adversity experiences | **1.37** | **0.21** | **2.02** | **0.04** | **1.01** | **1.85** |
| Total number of worries about adversity experiences | 1.16 | 0.17 | 0.97 | 0.33 | 0.86 | 1.55 |
| **Model 2** |  |  |  |  |  |  |
| Total number of adversity experiences | **1.42** | **0.22** | **2.25** | **0.02** | **1.05** | **1.94** |
| Total number of worries about adversity experiences | 1.13 | 0.16 | 0.82 | 0.41 | 0.85 | 1.50 |
| **Model 3** |  |  |  |  |  |  |
| Total number of adversity experiences | 1.20 | 0.21 | 1.06 | 0.29 | 0.86 | 1.69 |
| Total number of worries about adversity experiences | **1.47** | **0.26** | **2.17** | **0.03** | **1.04** | **2.08** |
| **Model 4** |  |  |  |  |  |  |
| Total number of adversity experiences | 1.23 | 0.23 | 1.11 | 0.27 | 0.85 | 1.76 |
| Total number of worries about adversity experiences | **1.46** | **0.27** | **2.06** | **0.04** | **1.02** | **2.09** |

Note. Data were weighted to the proportions of gender, age, ethnicity, country, and education obtained from the Office for National Statistics. Model 1 included only adversity experiences and worries about adversity experiences (in the same model), Model 2 additionally adjusted for COVID-19 infection variables, Model 3 additionally adjusted for socio-demographic characteristics, and Model 4 additionally adjusted for health-related factors.

**Table 7.** Logistic regressions predicting the development of difficulty with self-care from adversity experiences and worries about adversity experiences (N = 318), weighted

|  | Difficulty with self-care |  |  |  |  |  |
| --- | --- | --- | --- | --- | --- | --- |
|  | OR | SE | T | P | 95% CI | 95% CI |
| **Model 1** |  |  |  |  |  |  |
| Total number of adversity experiences | 1.08 | 0.13 | 0.69 | 0.49 | 0.86 | 1.36 |
| Total number of worries about adversity experiences | **1.49** | **0.28** | **2.17** | **0.03** | **1.04** | **2.15** |
| **Model 2** |  |  |  |  |  |  |
| Total number of adversity experiences | 1.05 | 0.12 | 0.44 | 0.66 | 0.84 | 1.32 |
| Total number of worries about adversity experiences | **1.45** | **0.25** | **2.18** | **0.03** | **1.04** | **2.03** |
| **Model 3** |  |  |  |  |  |  |
| Total number of adversity experiences | 1.05 | 0.14 | 0.35 | 0.73 | 0.81 | 1.36 |
| Total number of worries about adversity experiences | 1.32 | 0.22 | 1.67 | 0.10 | 0.95 | 1.84 |
| **Model 4** |  |  |  |  |  |  |
| Total number of adversity experiences | 1.06 | 0.15 | 0.43 | 0.67 | 0.80 | 1.41 |
| Total number of worries about adversity experiences | 1.30 | 0.22 | 1.55 | 0.12 | 0.93 | 1.81 |

Note. Data were weighted to the proportions of gender, age, ethnicity, country, and education obtained from the Office for National Statistics. Model 1 included only adversity experiences and worries about adversity experiences (in the same model), Model 2 additionally adjusted for COVID-19 infection variables, Model 3 additionally adjusted for socio-demographic characteristics, and Model 4 additionally adjusted for health-related factors.

### Sensitivity analyses

Results from sensitivity analyses which included people who were unsure whether they had had long COVID indicated similar findings for all analyses (Supplemental Tables S12). However, in the sensitivity analysis using COVID-19 symptom duration of more than four weeks as the outcome variable, the number of worries about adversity experiences was not associated with long COVID (OR: 1.14; 95% CI: 0.92 to 1.41) in the full model (Supplemental Table S13). There was, however, still evidence for an association of the lowest SEP category and the number of worries about adversity experiences with long COVID (Supplemental Table S14).

## Discussion

In this longitudinal study of UK adults, we explored the relationship between stress in the month prior to infection with COVID-19 and risk of developing long COVID and three long COVID symptoms. In our sample, one in five (20%) had either been diagnosed with or believed themselves to have had long COVID, which is similar to the 22% with any symptom five weeks after testing positive among Coronavirus Infection Survey respondents.^9^ We found that worries about adversity experiences in the month prior to infection were associated with a higher odds of developing long COVID. However, actual adversity experiences were not associated with increased odds of long COVID, although for every additional adversity people were worried about, there was 1.25 increased odds of long COVID. Once the SEP index was considered, those in the lowest SEP group were twice as likely as those in the highest to have developed long COVID, but there was no evidence for an interaction between the SEP index and the exposure variables. For individuals who had developed long COVID, there was also evidence that experiencing more worries about adversity experiences in the month prior to the infection increased the likelihood of difficulty with cognition (e.g., difficulty remembering or concentrating), but not with mobility or self-care.

The long COVID group had experienced more adversities in the month prior to becoming infected with COVID-19, including financial adversities, psychological or physical abuse, and were more likely to have not been able to access essentials such as food or medicine. They were also more worried about these adversities occurring than the group without long COVID. However, it was worrying about a range of adversity experiences in the month before being infected with COVID-19 that increased the likelihood of developing self-reported long COVID and difficulty with cognition. Other research using a sample of adults from the same dataset as the current study has found that worrying about experiencing adversities can be just as detrimental to mental health and sleep as experiencing those adversities.^17,45^ But it is notable that we did not find the same association for experiencing adversities. In the current study, it could be that there was a delay in the stress response associated with actual adversities such as losing one’s job,^46^ in particular as the impact in cuts in income set in^17^ and with repeated job search failures.^47^ Additionally, theories of stress and health posit that the impact of adversities depends on how well individuals are able to cope with stress.^48^ So the impact of adversities experienced during the pandemic, including falling ill with COVID-19, may have therefore been attenuated in individuals with more coping resources.

Our findings are relevant in our understanding of the biological mechanisms of long COVID. Whilst the aetiology underlying long COVID is complex and multi-factorial, two proposed biological mechanisms linking COVID-19 infection with long COVID are via immune system dysregulation and the reactivation of dormant viral fragments under stressful conditions.^49^ It is well known that stress leads to chronic inflammation,^50^ which in turn contributes to the burden of most preventable disease.^33^ Inflammation from stress prior to infection with COVID-19 may therefore play a key role in persistent symptoms/long COVID by enhancing the inflammatory response to COVID-19 infection. Therefore, it is plausible that immune system dysregulation due to stress before infection with COVID-19 may explain some of the relationship between COVID-19 and future impairment. Additionally, recent research has shown that individuals who develop long COVID experience a greater increase in anxiety and depressive symptoms in the immediate stage following infection, and can have slightly elevated anxiety symptoms in the weeks immediately preceding infection. The findings presented here help to explain why these anxiety symptoms may be elevated, by highlighting a relationship between stress and long COVID development, which may also help to explain the greater and persistent increase in anxiety in long COVID compared to short COVID patients post infection.

Our study also focused on the impact of the chronic stress of low SEP. The pandemic has imposed numerous financial and social stressors to many in the population, but not everyone has been affected equally. Lower SEP is linked to more severe COVID-19 infection^51^ and COVID-19 mortality.^52^ The association between SEP and chronic inflammation has been found consistently. People with lower incomes have not only been more adversely affected by adversity during the pandemic^54^ but were also more likely to face adversities before the pandemic, which may then result in heightened inflammation.^55,56^ Stress before infection with COVID-19 may lead to more severe infection,^57^ which has been shown to increase the risk for long COVID.^6,8,10^ In the current study, people with low household income and who had lower levels of education were more likely to develop long COVID. This echoes some previous research, which has found that living in a deprived area to be associated with long COVID,^9,15^ but contradicts other survey findings, which have not found links between self-reported income or education level and long COVID.^14,34^ However, there was no evidence of an interaction between adversity experiences and worries about adversity experiences and SEP. Therefore, it appears that the additional experience of adversities and related worries did not further increase the risk. It is notable that although previous research did suggest a slightly stronger association between adversity worries and experiences and negative effects on mental health in low SEP groups, this relationship was only very small.^17^ In considering the implications of these findings, funding for enhanced services for general practices in England to manage long COVID is distributed through registered number of patients, and does not consider factors influencing the prevalence of COVID-19 infection or long COVID, which may further increase inequalities.^58^ Given the increased risk of long COVID amongst individuals from low SEP backgrounds, more attention is arguably needed to ensure there are adequate resources to support these individuals, especially in areas of higher deprivation where funding allocation per long COVID patient may consequently be smaller.

We also focused on the relationship between low SEP, adversities, and specific symptoms of long COVID. Amongst those with self-reported long COVID in our study, over half reported having had difficulty with cognition (62%), which is higher than those reported by systematic reviews (e.g., 15%-26%^11,59^ and from the Coronavirus Infection Survey (25%).^9^ Having experienced a greater number of adversity experiences in this period increased the risk for difficulties with cognition (e.g., difficulty remembering or concentrating), but not for the other two long COVID symptoms measured (difficulties with mobility and self-care). ‘Brain fog’, or cognitive difficulties, is among the most commonly reported ongoing symptoms of COVID-19 and may persist even after other symptoms have cleared. For example, a study of over 80k individuals in the UK early in the pandemic found that even after recovery from COVID-19, significant cognitive impairment as evidenced by test scores remained, relative to controls who had not been infected with COVID-19.^60^ COVID-19 infection is also associated with subsequent mental health consequences, possibly as a result of penetration of the virus into the brain.^61^ More research on the psychosocial risk factors and biological mechanisms underpinning specific long COVID symptoms will facilitate the development of more effective treatments.

There are a number of strengths to the current study, including a large heterogenous sample which is well-stratified across major demographic groups. We also included a wide range of control variables, which, like our exposures, were measured prior to the outcome variables. However, the current study also has several limitations. First, systematic reviews have reported that there are a large number of different symptoms associated with long COVID,^2,11^ but we assessed a limited number of long COVID symptoms, and did not ask about fatigue, which is the most commonly reported long COVID symptoms.^9,10,12^ However, it is possible that participants interpreted our question on difficulties with mobility (e.g., walking or climbing steps) to mean difficulty carrying out these activities due to fatigue. Second, due to insufficient numbers, we were not able to include vaccination status at the time of infection as a covariate, which may decrease the likelihood of developing long COVID.^62,63^ A third limitation is the self-reported nature of our long COVID variable; participants were left to decide for themselves whether they believed themselves to have or have had long COVID. Although having been formally diagnosed with long COVID was a response option to this question, only around 1 in 5 (19%) had been diagnosed. The sample was also insufficiently powered to use symptom duration as our main outcome variable, as fewer than half who self-reported long COVID said their symptoms had lasted at least four weeks. To address how this may have influenced results, we conducted analyses comparing people whose symptoms had lasted fewer than four weeks with those whose symptoms lasted longer. Using this criterion, worries about adversity experiences predicted the outcome when adjusting for COVID-19 infection variables, but not once socio-demographics and health-related factors were included in the model. This difference in findings could have been due there being substantially fewer people in the long COVID case group when using this definition. Finally, the sample was insufficiently powered to detect the extent of difficulty participants had had with each of the three long COVID symptoms.

This work adds to the paucity of research on factors predictive of long COVID which were present before infection. Our longitudinal findings highlight the role of stress (both chronic stress such as low SEP and novel pandemic-related adversities and worries) as upstream determinants of long COVID and of cognitive difficulties specifically and point to the importance of policies which reassure people in times of social and financial uncertainty. It is clear that a substantial proportion of the population is at risk for ongoing symptoms after infection. Long COVID represents an emerging public health issue, and our findings point to stress preceding COVID-19 infection as a risk factor. Although free COVID-19 testing and financial support for people with COVID-19 was discontinued in England from 1 April 2022, the persistence of infection rates could lead to further disability, and a continued impact on individuals, their families, health services, and society. This highlights the importance of addressing novel and chronic stressors during pandemics both as a way of reducing adverse psychological effects but also of reducing the risk of long-term debilitating disease.

## Data Availability

The UCL COVID-19 Social Study documentation and codebook are available for download at https://osf.io/jm8ra/. Statistical code is available upon request from Elise Paul.

## Contributors

DF conceptualised and designed the study. DF also acquired funding, led the investigation, provided oversight on the methodology, administered the project, provided software and other resources, and supervised the project. Data were curated, validated, and formally analysed by EP. EP created visualisations, wrote the original manuscript draft with input from all authors. All authors reviewed and edited the manuscript. All authors approved the final version of the manuscript and had full access to and verified the data.

## Acknowledgements

The researchers are grateful for the support of a number of organisations with their recruitment efforts including: the UKRI Mental Health Networks, Find Out Now, UCL BioResource, SEO Works, FieldworkHub, and Optimal Workshop.

## Ethics approval and consent to participate

Ethical approval for the COVID-19 Social Study was granted by the UCL Ethics Committee. All participants provided fully informed consent and the study is GDPR compliant.

## Supplemental Materials

**Table S1.** Pattern of missing data in study sample (N = 1,966)

| Variable | Proportion with missing data |
| --- | --- |
| Household income | 7.94% |
| Employment status | 0% |
| Key worker status | 0% |
| Household crowding status | 0% |
| Living arrangement | 0% |
| Live in a rural area | 0% |
| COVID-19 infection severity | 0.81% |
| Dominant strain in the UK at time of COVID-19 infection | 0% |
| Long-term physical health condition | 0% |
| Long-term mental health condition | 0% |

**Table S2.** Complete case analysis: logistic regressions predicting the development of long COVID from adversity experiences and worries about adversity experiences (N = 1,520), weighted

|  | Self-reported long COVID |  |  |  |  |  |
| --- | --- | --- | --- | --- | --- | --- |
|  | OR | SE | T | P | 95% CI | 95% CI |
| **Model 1** |  |  |  |  |  |  |
| Total number of adversity experiences | 1.06 | 0.09 | 0.66 | 0.51 | 0.90 | 1.24 |
| Total number of worries about adversity experiences | **1.34** | **0.12** | **3.17** | **<0.001** | **1.12** | **1.60** |
| **Model 2** |  |  |  |  |  |  |
| Total number of adversity experiences | 1.04 | 0.09 | 0.53 | 0.60 | 0.89 | 1.23 |
| Total number of worries about adversity experiences | **1.29** | **0.12** | **2.81** | **<0.001** | **1.08** | **1.54** |
| **Model 3** |  |  |  |  |  |  |
| Total number of adversity experiences | 1.05 | 0.09 | 0.49 | 0.62 | 0.88 | 1.24 |
| Total number of worries about adversity experiences | **1.30** | **0.14** | **2.47** | **0.01** | **1.06** | **1.59** |
| **Model 4** |  |  |  |  |  |  |
| Total number of adversity experiences | 1.05 | 0.09 | 0.51 | 0.61 | 0.88 | 1.25 |
| Total number of worries about adversity experiences | **1.27** | **0.14** | **2.21** | **0.03** | **1.03** | **1.57** |

Note. Data were weighted to the proportions of gender, age, ethnicity, country, and education obtained from the Office for National Statistics. Model 1 included only adversity experiences and worries about adversity experiences (in the same model), Model 2 additionally adjusted for COVID-19 infection variables, Model 3 additionally adjusted for socio-demographic characteristics, and Model 4 additionally adjusted for health-related factors.

**Table S3.** Complete case analysis: logistic regressions predicting the development of long COVID from adversity experiences and worries about adversity experiences in interaction with low SEP (N = 1,520), weighted

|  |  | Self-reported long COVID |  |  |  |  |  |
| --- | --- | --- | --- | --- | --- | --- | --- |
|  |  | OR | SE | T | P | 95% CI | 95% CI |
| **Model 1** |  |  |  |  |  |  |  |
| Low SEP index (ref 0) |  |  |  |  |  |  |  |
|  | 1 | 0.94 | 0.25 | -0.24 | 0.81 | 0.56 | 1.57 |
|  | 2+ | **1.99** | **0.41** | **3.31** | **<0.001** | **1.32** | **2.99** |
| Total number of adversity experiences |  | 1.13 | 0.16 | 0.89 | 0.37 | 0.86 | 1.49 |
| Interaction with SEP |  |  |  |  |  |  |  |
|  | 1 | 0.94 | 0.21 | -0.30 | 0.77 | 0.60 | 1.45 |
|  | 2+ | 0.90 | 0.16 | -0.58 | 0.56 | 0.63 | 1.28 |
| Total number of worries about adversity experiences |  | **1.42** | **0.20** | **2.53** | **0.01** | **1.08** | **1.86** |
| Interaction with SEP |  |  |  |  |  |  |  |
|  | 1 | 0.80 | 0.15 | -1.18 | 0.24 | 0.55 | 1.16 |
|  | 2+ | 0.99 | 0.20 | -0.06 | 0.95 | 0.66 | 1.47 |
| **Model 2** |  |  |  |  |  |  |  |
| Low SEP index (ref 0) |  |  |  |  |  |  |  |
|  | 1 | 0.93 | 0.24 | -0.30 | 0.76 | 0.56 | 1.53 |
|  | 2+ | **2.03** | **0.44** | **3.24** | **<0.001** | **1.32** | **3.12** |
| Total number of adversity experiences |  | 1.12 | 0.16 | 0.81 | 0.42 | 0.85 | 1.48 |
| Interaction with SEP |  |  |  |  |  |  |  |
|  | 1 | 0.91 | 0.20 | -0.43 | 0.67 | 0.59 | 1.41 |
|  | 2+ | 0.91 | 0.17 | -0.51 | 0.61 | 0.64 | 1.30 |
| Total number of worries about adversity experiences |  | **1.51** | **0.21** | **2.89** | **<0.001** | **1.14** | **1.99** |
| Interaction with SEP |  |  |  |  |  |  |  |
|  | 1 | 0.69 | 0.15 | -1.74 | 0.08 | 0.46 | 1.05 |
|  | 2+ | 0.89 | 0.19 | -0.57 | 0.57 | 0.59 | 1.34 |
| **Model 3** |  |  |  |  |  |  |  |
| Low SEP index (ref 0) |  |  |  |  |  |  |  |
|  | 1 | 0.85 | 0.21 | -0.64 | 0.52 | 0.52 | 1.39 |
|  | 2+ | **2.30** | **0.58** | **3.30** | **<0.001** | **1.40** | **3.78** |
| Total number of adversity experiences |  | 1.10 | 0.18 | 0.59 | 0.56 | 0.80 | 1.50 |
| Interaction with SEP |  |  |  |  |  |  |  |
|  | 1 | 0.93 | 0.23 | -0.31 | 0.76 | 0.58 | 1.49 |
|  | 2+ | 0.96 | 0.19 | -0.18 | 0.85 | 0.65 | 1.42 |
| Total number of worries about adversity experiences |  | **1.55** | **0.24** | **2.76** | **0.01** | **1.14** | **2.11** |
| Interaction with SEP |  |  |  |  |  |  |  |
|  | 1 | 0.67 | 0.15 | -1.74 | 0.08 | 0.43 | 1.05 |
|  | 2+ | 0.83 | 0.19 | -0.80 | 0.42 | 0.53 | 1.31 |
| **Model 4** |  |  |  |  |  |  |  |
| Low SEP index (ref 0) |  |  |  |  |  |  |  |
|  | 1 | 0.83 | 0.21 | -0.74 | 0.46 | 0.50 | 1.37 |
|  | 2+ | **2.15** | **0.55** | **2.98** | **<0.001** | **1.30** | **3.56** |
| Total number of adversity experiences |  | 1.10 | 0.18 | 0.58 | 0.56 | 0.80 | 1.51 |
| Interaction with SEP |  |  |  |  |  |  |  |
|  | 1 | 0.94 | 0.23 | -0.25 | 0.80 | 0.59 | 1.51 |
|  | 2+ | 0.96 | 0.19 | -0.19 | 0.85 | 0.65 | 1.42 |
| Total number of worries about adversity experiences |  | **1.55** | **0.25** | **2.72** | **0.01** | **1.13** | **2.12** |
| Interaction with SEP |  |  |  |  |  |  |  |
|  | 1 | 0.65 | 0.15 | -1.85 | 0.06 | 0.42 | 1.03 |
|  | 2+ | 0.82 | 0.19 | -0.84 | 0.40 | 0.52 | 1.30 |

Note. Data were weighted to the proportions of gender, age, ethnicity, country, and education obtained from the Office for National Statistics. Model 1 included only adversity experiences and worries about adversity experiences (in the same model), Model 2 additionally adjusted for COVID-19 infection variables, Model 3 additionally adjusted for socio-demographic characteristics, and Model 4 additionally adjusted for health-related factors.

**Table S4.** Complete case analysis: logistic regressions predicting the development of difficulty with mobility from adversity experiences and worries about adversity experiences (N = 290), weighted

|  | Difficulty with mobility |  |  |  |  |  |
| --- | --- | --- | --- | --- | --- | --- |
|  | OR | SE | T | P | 95% CI | 95% CI |
| **Model 1** |  |  |  |  |  |  |
| Total number of adversity experiences | 1.29 | 0.19 | 1.77 | 0.08 | 0.97 | 1.72 |
| Total number of worries about adversity experiences | 0.93 | 0.16 | -0.40 | 0.69 | 0.67 | 1.30 |
| **Model 2** |  |  |  |  |  |  |
| Total number of adversity experiences | **1.33** | **0.19** | **1.96** | **0.05** | **1.00** | **1.76** |
| Total number of worries about adversity experiences | 0.93 | 0.15 | -0.46 | 0.65 | 0.67 | 1.28 |
| **Model 3** |  |  |  |  |  |  |
| Total number of adversity experiences | **1.36** | **0.21** | **2.00** | **0.05** | **1.01** | **1.84** |
| Total number of worries about adversity experiences | 0.93 | 0.16 | -0.39 | 0.70 | 0.66 | 1.32 |
| **Model 4** |  |  |  |  |  |  |
| Total number of adversity experiences | 1.38 | 0.23 | 1.92 | 0.06 | 0.99 | 1.93 |
| Total number of worries about adversity experiences | 0.96 | 0.18 | -0.23 | 0.82 | 0.66 | 1.38 |

Note. Data were weighted to the proportions of gender, age, ethnicity, country, and education obtained from the Office for National Statistics. Model 1 included only adversity experiences and worries about adversity experiences (in the same model), Model 2 additionally adjusted for COVID-19 infection variables, Model 3 additionally adjusted for socio-demographic characteristics, and Model 4 additionally adjusted for health-related factors.

**Table S5.** Complete case analysis: logistic regressions predicting the development of difficulty with cognition from adversity experiences and worries about adversity experiences (N = 290), weighted

|  | Difficulty with cognition |  |  |  |  |  |
| --- | --- | --- | --- | --- | --- | --- |
|  | OR | SE | T | P | 95% CI | 95% CI |
| **Model 1** |  |  |  |  |  |  |
| Total number of adversity experiences | **1.39** | **0.23** | **2.04** | **0.04** | **1.01** | **1.92** |
| Total number of worries about adversity experiences | 1.06 | 0.19 | 0.35 | 0.73 | 0.75 | 1.51 |
| **Model 2** |  |  |  |  |  |  |
| Total number of adversity experiences | **1.43** | **0.23** | **2.19** | **0.03** | **1.04** | **1.97** |
| Total number of worries about adversity experiences | 1.06 | 0.18 | 0.36 | 0.72 | 0.76 | 1.49 |
| **Model 3** |  |  |  |  |  |  |
| Total number of adversity experiences | 1.14 | 0.20 | 0.73 | 0.47 | 0.81 | 1.60 |
| Total number of worries about adversity experiences | 1.51 | 0.34 | 1.85 | 0.06 | 0.98 | 2.33 |
| **Model 4** |  |  |  |  |  |  |
| Total number of adversity experiences | 1.17 | 0.22 | 0.81 | 0.42 | 0.80 | 1.69 |
| Total number of worries about adversity experiences | 1.50 | 0.36 | 1.70 | 0.09 | 0.94 | 2.40 |

Note. Data were weighted to the proportions of gender, age, ethnicity, country, and education obtained from the Office for National Statistics. Model 1 included only adversity experiences and worries about adversity experiences (in the same model), Model 2 additionally adjusted for COVID-19 infection variables, Model 3 additionally adjusted for socio-demographic characteristics, and Model 4 additionally adjusted for health-related factors.

**Table S6.** Complete case analysis: logistic regressions predicting the development of difficulty with self-care from adversity experiences and worries about adversity experiences (N = 290), weighted

|  | Difficulty with self-care |  |  |  |  |  |
| --- | --- | --- | --- | --- | --- | --- |
|  | OR | SE | T | P | 95% CI | 95% CI |
| **Model 1** |  |  |  |  |  |  |
| Total number of adversity experiences | 1.20 | 0.13 | 1.74 | 0.08 | 0.98 | 1.47 |
| Total number of worries about adversity experiences | 1.30 | 0.27 | 1.26 | 0.21 | 0.87 | 1.94 |
| **Model 2** |  |  |  |  |  |  |
| Total number of adversity experiences | 1.16 | 0.13 | 1.34 | 0.18 | 0.93 | 1.44 |
| Total number of worries about adversity experiences | 1.28 | 0.26 | 1.23 | 0.22 | 0.86 | 1.89 |
| **Model 3** |  |  |  |  |  |  |
| Total number of adversity experiences | 1.14 | 0.15 | 0.96 | 0.34 | 0.87 | 1.48 |
| Total number of worries about adversity experiences | 1.20 | 0.22 | 1.03 | 0.30 | 0.85 | 1.71 |
| **Model 4** |  |  |  |  |  |  |
| Total number of adversity experiences | 1.17 | 0.16 | 1.13 | 0.26 | 0.89 | 1.53 |
| Total number of worries about adversity experiences | 1.17 | 0.21 | 0.88 | 0.38 | 0.82 | 1.66 |

Note. Data were weighted to the proportions of gender, age, ethnicity, country, and education obtained from the Office for National Statistics. Model 1 included only adversity experiences and worries about adversity experiences (in the same model), Model 2 additionally adjusted for COVID-19 infection variables, Model 3 additionally adjusted for socio-demographic characteristics, and Model 4 additionally adjusted for health-related factors.

**Table S7.**
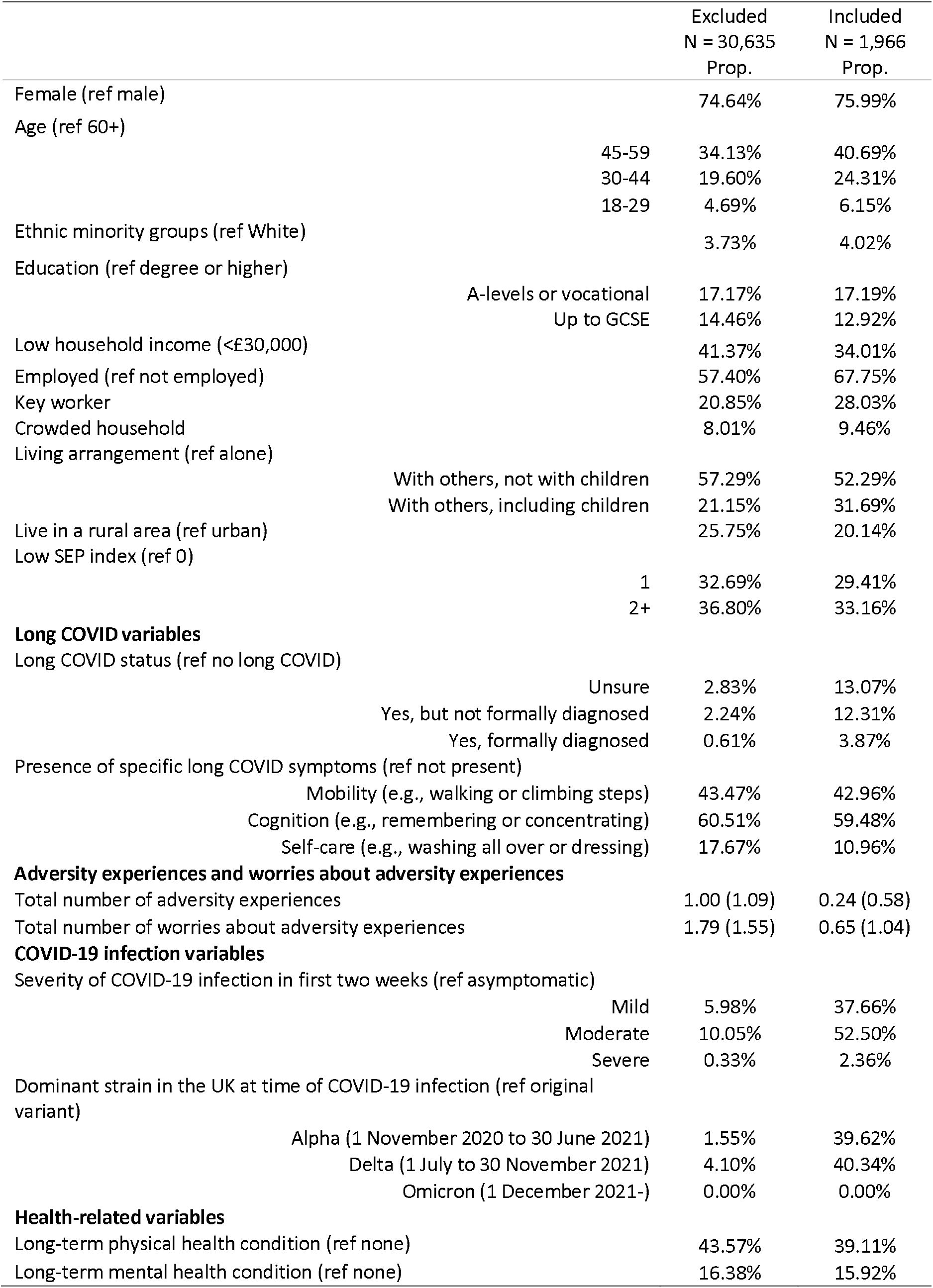
Characteristics of excluded and included participants, unweighted

**Table S8.**
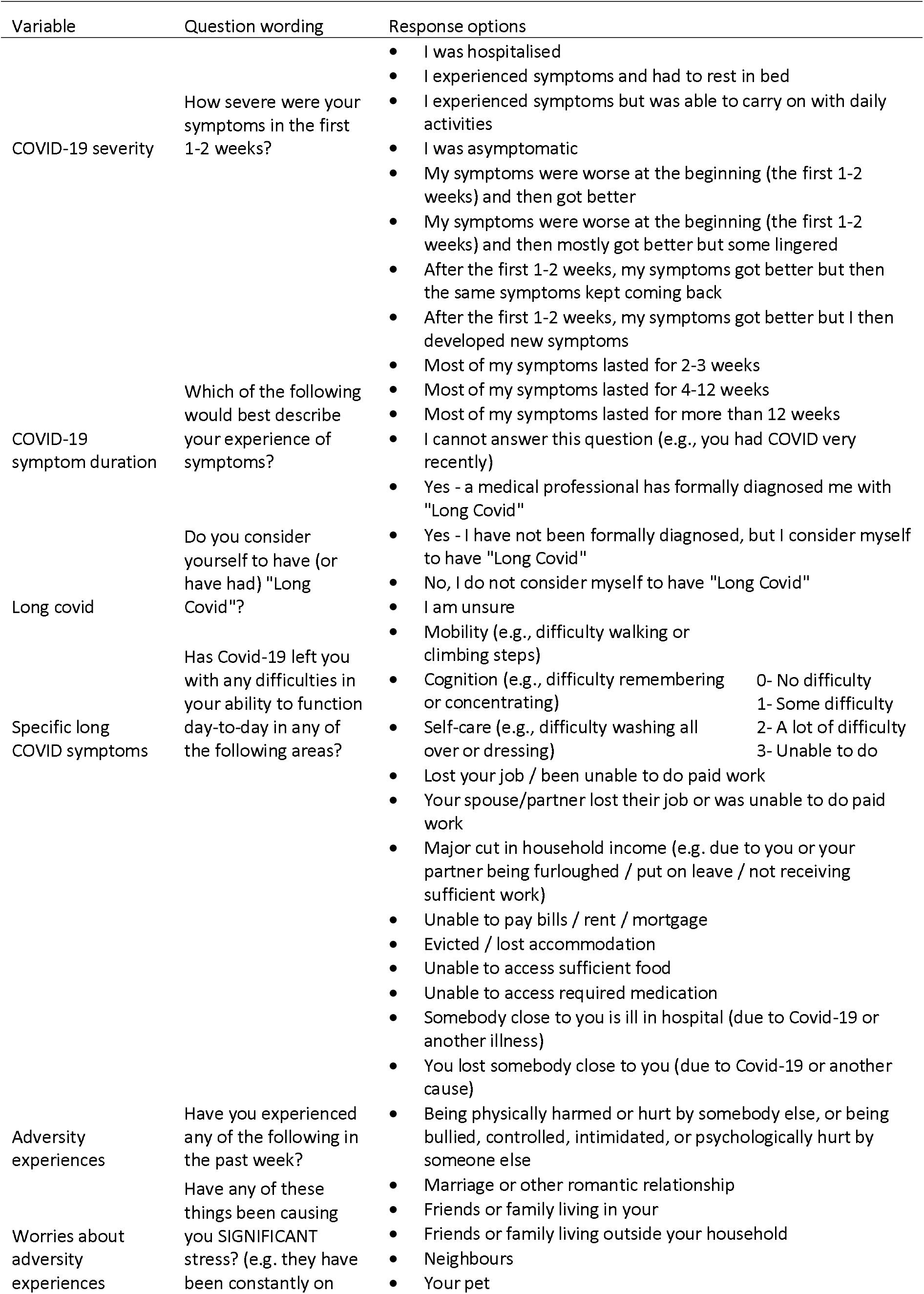
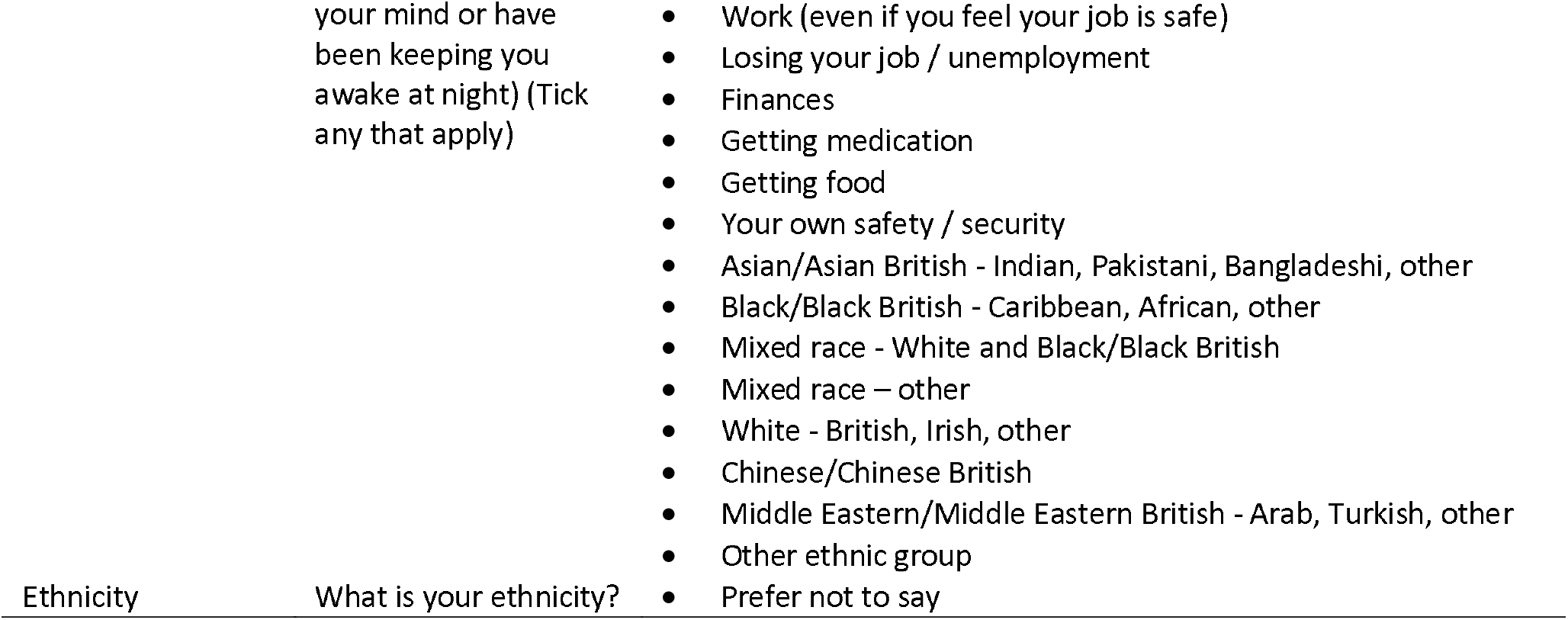
Wording of study developed items

**Table S9.**
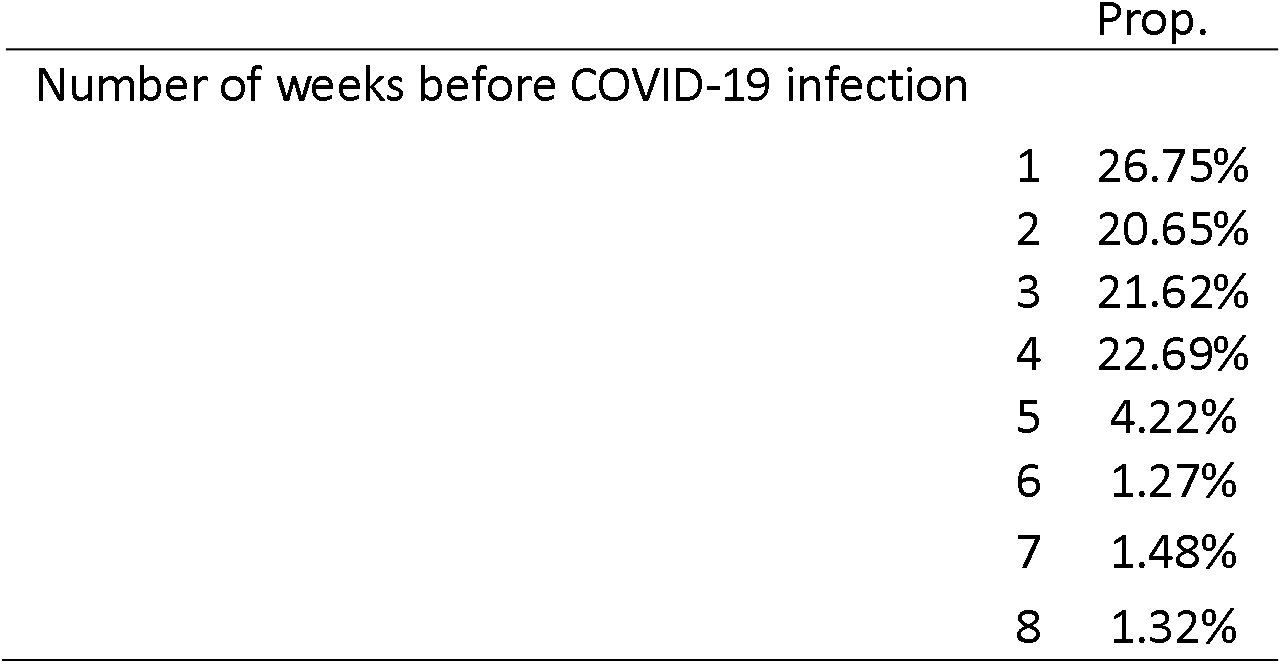
Proportion of participants with data for week closest to COVID-19 infection (N = 1,966)

**Table S10.** Weighted and unweighted characteristics of the sample (N = 1,966)

|  |  | Unweighted<br>Prop. | Weighted<br>Prop. |
| --- | --- | --- | --- |
| **Socio-demographics** |  |  |  |
| Female (ref male) |  | 75.99% | 52.48% |
| Age (ref 60+) |  |  |  |
|  | 45-59 | 40.69% | 32.15% |
|  | 30-44 | 24.31% | 20.55% |
|  | 18-29 | 6.15% | 10.97% |
| Ethnic minority groups (ref White) |  | 4.02% | 7.17% |
| Education (ref degree or higher) |  |  |  |
|  | A-levels or vocational | 17.19% | 32.74% |
|  | Up to GCSE | 12.92% | 29.72% |
| Low household income (<£30,000) |  | 34.01% | 42.33% |
| Employed (ref not employed) |  | 67.75% | 59.94% |
| Key worker |  | 28.03% | 25.43% |
| Crowded household |  | 9.46% | 11.84% |
| Living arrangement (ref alone) |  |  |  |
|  | With others, not with children | 52.29% | 54.77% |
|  | With others, including children | 31.69% | 28.12% |
| Live in a rural area (ref urban) |  | 20.14% | 19.49% |
| Low SEP index (ref 0) |  |  |  |
|  | 1 | 29.41% | 29.77% |
|  | 2+ | 33.16% | 46.98% |
| **Long COVID variables** |  |  |  |
| Long COVID status (ref no long COVID) |  |  |  |
|  | Unsure | 13.07% | 12.41% |
|  | Yes, but not formally diagnosed | 12.31% | 16.34% |
|  | Yes, formally diagnosed | 3.87% | 3.76% |
| Presence of specific long COVID symptoms (ref not present) |  |  |  |
|  | Mobility (e.g., walking or climbing steps) | 42.96% | 44.06% |
|  | Cognition (e.g., remembering or concentrating) | 59.48% | 54.09% |
|  | Self-care (e.g., washing all over or dressing) | 10.96% | 11.96% |
| **Adversity experiences and worries about adversity experiences** |  |  |  |
| Total number of adversity experiences |  | 0.24 (0.58) | 0.23 (0.55) |
| Total number of worries about adversity experiences |  | 0.65 (1.04) | 0.64 (1.09) |
| **COVID-19 infection variables** |  |  |  |
| COVID-19 infection severity (ref asymptomatic) |  |  |  |
|  | Mild | 37.66% | 40.14% |
|  | Moderate | 52.50% | 49.45% |
|  | Severe | 2.36% | 2.91% |
| Dominant strain in the UK at time of COVID-19 infection (ref original variant) |  |  |  |
|  | Alpha (1 November 2020 to 30 June 2021) | 39.62% | 36.67% |
|  | Delta (1 July to 30 November 2021) | 40.34% | 42.55% |
|  | Omicron (1 December 2021-) | 0.00% | 0.00% |
| **Health-related variables** |  |  |  |
| Long-term physical health condition (ref none) |  | 39.11% | 44.02% |
| Long-term mental health condition (ref none) |  | 15.92% | 15.62% |

Note. Data in the weighted sample were weighted to the proportions of gender, age, ethnicity, country, and education obtained from the Office for National Statistics. GCSE refers to General Certificate of Secondary Education.

**Table S11.** Sensitivity analysis: logistic regressions predicting self-reported long COVID adversity experiences and worries about adversity experiences, with participants who were ‘unsure’ whether they had had long COVID in the case group (N= 1,966), weighted

|  | Self-reported long COVID |  |  |  |  |  |
| --- | --- | --- | --- | --- | --- | --- |
|  | OR | SE | T | P | 95% CI | 95% CI |
| **Model 1** |  |  |  |  |  |  |
| Total number of adversity experiences | 1.03 | 0.07 | 0.43 | 0.67 | 0.90 | 1.18 |
| Total number of worries about adversity experiences | 1.30 | 0.10 | 3.46 | **<0.001** | 1.12 | 1.51 |
| **Model 2** |  |  |  |  |  |  |
| Total number of adversity experiences | 1.01 | 0.07 | 0.18 | 0.86 | 0.88 | 1.16 |
| Total number of worries about adversity experiences | 1.25 | 0.09 | 3.00 | **<0.001** | 1.08 | 1.45 |
| **Model 3** |  |  |  |  |  |  |
| Total number of adversity experiences | 1.02 | 0.07 | 0.22 | 0.82 | 0.88 | 1.17 |
| Total number of worries about adversity experiences | **1.23** | **0.10** | **2.61** | **0.01** | **1.05** | **1.44** |
| **Model 4** |  |  |  |  |  |  |
| Total number of adversity experiences | 1.02 | 0.07 | 0.25 | 0.80 | 0.88 | 1.17 |
| Total number of worries about adversity experiences | **1.22** | **0.10** | **2.48** | **0.01** | **1.04** | **1.43** |

Note. Data were weighted to the proportions of gender, age, ethnicity, country, and education obtained from the Office for National Statistics. Model 1 included only adversity experiences and worries about adversity experiences (in the same model), Model 2 additionally adjusted for COVID-19 infection variables, Model 3 additionally adjusted for socio-demographic characteristics, and Model 4 additionally adjusted for health-related factors.

**Table S12.**
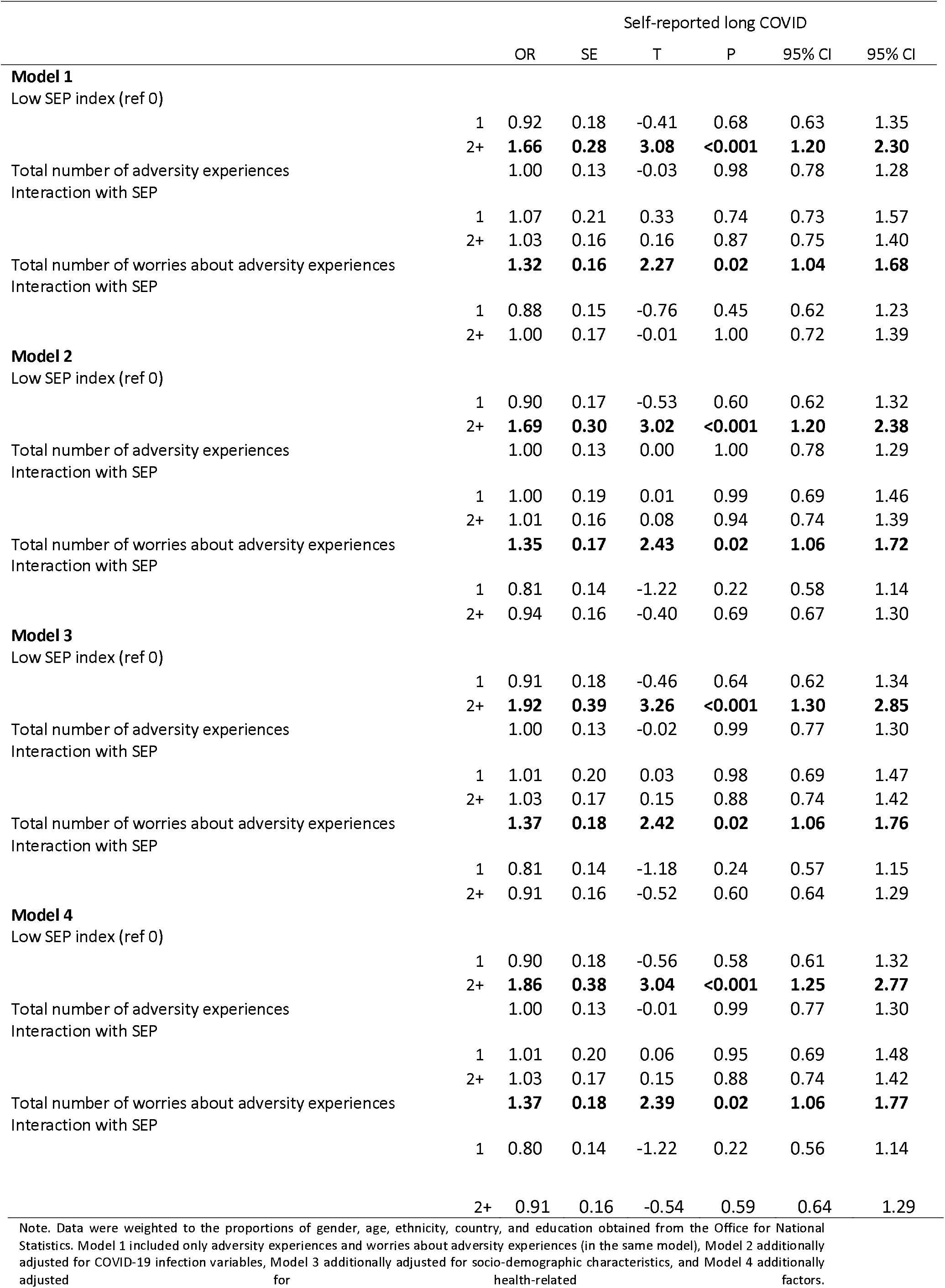
Sensitivity analysis: logistic regressions predicting self-reported long COVID adversity experiences and worries about adversity experiences, with participants who were ‘unsure’ whether they had had long COVID in the case group (N= 1,966), weighted

**Table S13.** Sensitivity analysis: logistic regressions predicting COVID-19 symptoms lasting four weeks or more from adversity experiences and worries about adversity experiences (N = 1,966), weighted

|  | COVID-19 symptoms of at least four weeks |  |  |  |  |  |
| --- | --- | --- | --- | --- | --- | --- |
|  | OR | SE | T | P | 95% CI | 95% CI |
| **Model 1** |  |  |  |  |  |  |
| Total number of adversity experiences | 0.98 | 0.10 | -0.21 | 0.84 | 0.79 | 1.21 |
| Total number of worries about adversity experiences | **1.27** | **0.12** | **2.51** | **0.01** | **1.05** | **1.52** |
| **Model 2** |  |  |  |  |  |  |
| Total number of adversity experiences | 0.97 | 0.11 | -0.29 | 0.77 | 0.78 | 1.20 |
| Total number of worries about adversity experiences | **1.23** | **0.12** | **2.14** | **0.03** | **1.02** | **1.48** |
| **Model 3** |  |  |  |  |  |  |
| Total number of adversity experiences | 0.97 | 0.10 | -0.31 | 0.76 | 0.78 | 1.20 |
| Total number of worries about adversity experiences | 1.16 | 0.12 | 1.39 | 0.16 | 0.94 | 1.43 |
| **Model 4** |  |  |  |  |  |  |
| Total number of adversity experiences | 0.96 | 0.11 | -0.37 | 0.71 | 0.77 | 1.19 |
| Total number of worries about adversity experiences | 1.14 | 0.12 | 1.19 | 0.23 | 0.92 | 1.41 |

Note. Data were weighted to the proportions of gender, age, ethnicity, country, and education obtained from the Office for National Statistics. Model 1 included only adversity experiences and worries about adversity experiences (in the same model), Model 2 additionally adjusted for COVID-19 infection variables, Model 3 additionally adjusted for socio-demographic characteristics, and Model 4 additionally adjusted for health-related factors.

**Table S14.** Sensitivity analysis: logistic regressions predicting COVID-19 symptoms lasting four weeks or more from adversity experiences and worries about adversity experiences in interaction with low SEP (N = 1,966), weighted

|  |  | Self-reported long COVID |  |  |  |  |  |
| --- | --- | --- | --- | --- | --- | --- | --- |
|  |  | OR | SE | T | P | 95% CI | 95% CI |
| **Model 1** |  |  |  |  |  |  |  |
| Low SEP index (ref 0) |  |  |  |  |  |  |  |
|  | 1 | 1.06 | 0.34 | 0.19 | 0.85 | 0.57 | 1.99 |
|  | 2+ | **2.54** | **0.67** | **3.55** | **<0.001** | **1.52** | **4.25** |
| Total number of adversity experiences |  | 1.23 | 0.20 | 1.29 | 0.20 | 0.90 | 1.70 |
| Interaction with SEP |  |  |  |  |  |  |  |
|  | 1 | 0.80 | 0.22 | -0.83 | 0.41 | 0.47 | 1.36 |
|  | 2+ | 0.74 | 0.17 | -1.34 | 0.18 | 0.48 | 1.15 |
| Total number of worries about adversity experiences |  | **1.52** | **0.25** | **2.53** | **0.01** | **1.10** | **2.10** |
| Interaction with SEP |  |  |  |  |  |  |  |
|  | 1 | 0.88 | 0.18 | -0.62 | 0.53 | 0.58 | 1.32 |
|  | 2+ | 0.77 | 0.16 | -1.25 | 0.21 | 0.51 | 1.16 |
| **Model 2** |  |  |  |  |  |  |  |
| Low SEP index (ref 0) |  |  |  |  |  |  |  |
|  | 1 | 1.03 | 0.33 | 0.08 | 0.93 | 0.55 | 1.91 |
|  | 2+ | **2.51** | **0.67** | **3.48** | **<0.001** | **1.50** | **4.22** |
| Total number of adversity experiences |  | 1.20 | 0.20 | 1.10 | 0.27 | 0.87 | 1.65 |
| Interaction with SEP |  |  |  |  |  |  |  |
|  | 1 | 0.78 | 0.21 | -0.91 | 0.36 | 0.46 | 1.32 |
|  | 2+ | 0.77 | 0.18 | -1.12 | 0.26 | 0.49 | 1.21 |
| Total number of worries about adversity experiences |  | **1.53** | **0.25** | **2.67** | **0.01** | **1.12** | **2.10** |
| Interaction with SEP |  |  |  |  |  |  |  |
|  | 1 | 0.85 | 0.18 | -0.80 | 0.42 | 0.56 | 1.27 |
|  | 2+ | 0.73 | 0.15 | -1.52 | 0.13 | 0.48 | 1.10 |
| **Model 3** |  |  |  |  |  |  |  |
| Low SEP index (ref 0) |  |  |  |  |  |  |  |
|  | 1 | 1.06 | 0.33 | 0.19 | 0.85 | 0.57 | 1.97 |
|  | 2+ | **2.74** | **0.83** | **3.35** | **<0.001** | **1.52** | **4.95** |
| Total number of adversity experiences |  | 1.16 | 0.20 | 0.85 | 0.40 | 0.83 | 1.62 |
| Interaction with SEP |  |  |  |  |  |  |  |
|  | 1 | 0.77 | 0.22 | -0.90 | 0.37 | 0.44 | 1.35 |
|  | 2+ | 0.79 | 0.19 | -0.99 | 0.32 | 0.50 | 1.25 |
| Total number of worries about adversity experiences |  | **1.51** | **0.24** | **2.55** | **0.01** | **1.10** | **2.07** |
| Interaction with SEP |  |  |  |  |  |  |  |
|  | 1 | 0.85 | 0.18 | -0.79 | 0.43 | 0.56 | 1.28 |
|  | 2+ | 0.73 | 0.16 | -1.46 | 0.15 | 0.48 | 1.12 |
| **Model 4** |  |  |  |  |  |  |  |
| Low SEP index (ref 0) |  |  |  |  |  |  |  |
|  | 1 | 1.03 | 0.32 | 0.10 | 0.92 | 0.56 | 1.90 |
|  | 2+ | **2.50** | **0.75** | **3.07** | **<0.001** | **1.39** | **4.50** |
| Total number of adversity experiences |  | 1.18 | 0.19 | 1.00 | 0.32 | 0.86 | 1.62 |
| Interaction with SEP |  |  |  |  |  |  |  |
|  | 1 | 0.77 | 0.22 | -0.92 | 0.36 | 0.45 | 1.34 |
|  | 2+ | 0.78 | 0.18 | -1.11 | 0.27 | 0.50 | 1.22 |
| Total number of worries about adversity experiences |  | **1.52** | **0.25** | **2.55** | **0.01** | **1.10** | **2.10** |
| Interaction with SEP |  |  |  |  |  |  |  |
|  | 1 | 0.82 | 0.17 | -0.96 | 0.34 | 0.54 | 1.24 |
|  | 2+ | 0.70 | 0.15 | -1.60 | 0.11 | 0.46 | 1.08 |

Note. Data were weighted to the proportions of gender, age, ethnicity, country, and education obtained from the Office for National Statistics. Model 1 included only adversity experiences and worries about adversity experiences (in the same model), Model 2 additionally
adjusted for COVID-19 infection variables, Model 3 additionally adjusted for socio-demographic characteristics, and Model 4 additionally adjusted for health-related factors.

